# Stroke Education in Student-Run Free Clinics: Identifying and Closing Knowledge Gaps in Underserved Populations

**DOI:** 10.64898/2026.01.16.26344279

**Authors:** Marcus Milani, Kiaya Johnston, Sophie Rewey, Brian Sick

## Abstract

**Background:** Stroke is a leading cause of death and disability, particularly in underserved populations with limited healthcare access, where poor health literacy and low stroke awareness contribute to delayed symptom recognition and worse outcomes. Student-run free clinics serve high-risk patients with hypertension, diabetes, and dyslipidemia, which are key stroke risk factors, yet stroke awareness remains inadequate. This study aims to identify stroke knowledge gaps and develop a culturally relevant educational intervention.

**Methods:** The three-phase study includes baseline surveys, educational material development, and post-education evaluation.

**Results:** After participation in stroke education, significant improvements in stroke sign recognition were observed, with Spanish speakers showing gains in knowledge about balance and vision loss, and English speakers in balance, vision loss, and face drooping (p <0.01). Emergency response knowledge improved less consistently; calling 911 significantly increased among English speakers (p <0.01) but decreased significantly (p <0.01) among Spanish speakers. The intervention effectively closed specific risk factor gaps, such as alcohol recognition among Spanish speakers (p <0.01).

**Conclusion:** By addressing knowledge gaps and empowering patients to act quickly, this intervention may reduce stroke-related morbidity and mortality in high-risk populations. The model could also serve as a framework for other student-run clinics to address health literacy gaps in underserved communities.

## Introduction

Stroke remains a leading cause of death and long-term disability worldwide, disproportionately affecting populations with limited access to healthcare resources.^1^ National surveys indicate that only about two-thirds of U.S. adults can correctly identify all major stroke warning signs, with significantly lower awareness among individuals with limited English proficiency.^2–4^ This disparity is pronounced in African American and Hispanic communities, who experience elevated stroke risk yet often have lower recognition of stroke symptoms and delayed access to treatments like thrombolytic therapy.^5^ Additionally, Hispanic individuals encounter greater challenges in stroke prevention due to lower health literacy and reduced awareness of acute interventions.^6,7^ These findings underscore the critical need for culturally tailored educational programs to improve stroke knowledge in vulnerable groups. However, many current stroke education tools remain inadequate, often due to poor accessibility, lack of interactivity, and failure to address the complex health literacy demands of stroke prevention and management.^8^

The educational gaps are exacerbated in low resource settings, where patients often face significant barriers to health information, increasing their risk of poor stroke outcomes.^9,10^ These populations frequently have a high prevalence of stroke risk factors, including hypertension, type II diabetes mellitus, and dyslipidemia, which further elevates their lifetime stroke risk.^11^

Student-run free clinics are well positioned to address this gap by delivering brief, no-cost preventative services tailored to the cultural and linguistic needs of underserved communities. These clinics often serve the uninsured, racial and ethnic minorities, immigrants, and non-English speakers, all populations with identified deficits in stroke health literacy.^12–17^ Given that stroke outcomes heavily depend on rapid recognition and treatment, targeted preemptive interventions in these high-risk populations are crucial.^18^ While community-based initiatives, such as school and community-based education campaigns, have shown promise,^19,20^ further research is needed to tailor stroke education for diverse populations with varying language, cultural, and health literacy needs.

This study assesses baseline stroke knowledge among patients at a student-run free clinic by identifying critical gaps in awareness that may contribute to delays in symptom recognition and treatment. By implementing and evaluating a tailored education intervention, this project aims to empower patients with the knowledge needed to recognize stroke symptoms early and seek timely medical care. Furthermore, this research may serve as a model for similar initiatives in other underserved settings, contributing to broader efforts to reduce stroke-related health disparities.

## Methods

### Participants and Recruitment

The study population consisted of patients attending the Phillips Neighborhood Clinic (PNC), a student-run, free clinic in Minneapolis, Minnesota. Inclusion criteria required participants to be 18 years of age or older and to have the capacity to provide informed consent. Exclusion criteria included: students or employees of the researchers, adults with diminished capacity to consent, and individuals approached during a stressful situation. Potential participants were initially screened for inclusion and exclusion criteria by PNC clinic volunteers at check-in. Patients expressing interest were provided with an information sheet, and final eligibility determination was conducted by trained research staff listed on the study protocol. This study was determined exempt from review by the University of Minnesota Institutional Review Board (STUDY00023210).

### Surveys

A researcher-developed survey was employed to collect data. See Appendix A for the survey instrument used in this investigation. Although several validated stroke knowledge assessment tools exist, such as the Stroke Knowledge Test and the Stroke Action Test, these instruments did not address key variables central to the present investigation. Specifically, existing tools typically rely on recognition of stroke symptoms and appropriate responses but omit critical sociodemographic and behavioral dimensions relevant to this study’s target population.

Our investigation aimed to explore factors beyond symptom knowledge alone, including country of origin, language preference, and gender identity (with inclusion of a non-binary response option). Additionally, the survey sought to assess knowledge of modifiable stroke risk factors, areas not included in all validated tools. For these reasons, a tailored researcher-developed instrument was designed to better capture both stroke literacy and the social determinants influencing stroke awareness in a linguistically and culturally diverse population.

### Baseline Data Collection

Baseline data were collected from a group of 62 (32 Spanish, 30 English) patients at the PNC. Eligible participants completed a paper-based survey designed to assess demographic characteristics and health literacy specific to stroke knowledge. This baseline pre-intervention data was used to inform the development of targeted educational materials for the intervention, while post-intervention data were analyzed to assess changes in response to the education provided. Spanish-language and English-language surveys were analyzed separately.

### Stroke Education Design

The educational intervention’s content was informed by broader literature and categories demonstrating the lowest baseline survey responses. The literature indicates that stroke outcomes are most effectively improved by increasing stroke sign and symptom awareness and time sensitivity in response.^10^ Literature also points to the utility of validated tools to teach this, especially the BE FAST/RÁPIDO model, known for its English–Spanish population utility.^29^ The model inherently emphasizes the urgency of time-sensitive response, its very acronyms (“BE FAST!” and “RÁPIDO!”) reinforce the need for immediate action. Baseline results, presented more fully in the Results section below, revealed markedly low recognition of vision loss as a symptom of stroke in both language groups and moderately low responses for balance, face drooping, speech problems, and limb or arm weakness. Additionally, both language groups demonstrated suboptimal recognition of the need to promptly seek healthcare or call 911 when stroke symptoms occur, reflecting a gap in time-sensitive response behaviors. Given the high percentage of patients from outside the US, as described in the Result section, educational materials needed to be culturally sensitive and cannot assume prior knowledge of calling emergency services. Centering the BEFAST/RÁPIDO model therefore appeared efficacious in reducing stroke knowledge gaps in the population, as supported by both the broader literature and baseline survey results.

Furthermore, the literature supports that recognizing modifiable risk factors for stroke can help motivate people to access primary care and overcome stroke knowledge gaps.^22^ Baseline survey results indicated low recognition of all modifiable risk factors, and especially low recognition of alcohol, diet, and stress. As such, the educational intervention also included all modifiable risk factors, with special emphasis on those with low baseline recognition. The average age of the pre-intervention group was 48.8 years and lack of participants without a history of stroke highlighted the importance of primary prevention.

Baseline surveys also demonstrated generally low recognition of stroke definitions (hemorrhagic vs ischemic). Given the low baseline knowledge, we sought to fill this gap in the surveyed population by including this topic; though only briefly, as limited evidence supports that such content directly improves stroke outcomes.

Educational materials included two parts: (1) an educational poster to be utilized in the clinic alongside a volunteer delivering a standardized stroke education talk, and (2) take-home brochures reiterating the material taught. Principles of Graphic Medicine were employed in the design of educational materials. Graphic medicine is defined as the intersection between the medium of comics and the discourse of healthcare.^23^ Graphic medicine combines text and art to clarify complex medical topics. In our educational materials, we used graphic medicine principles to explain the pathophysiology, signs, and risk factors of stroke to a patient population. The use of visuals allows graphic medicine to transcend language and education barriers, making the tool an affordable and accessible option in patient education.^24^ Educational tools were validated by stroke professionals. All content was offered in both English and Spanish, with Spanish content translated by native speakers shown in Appendix B and C.

### Post-Intervention Data Collection

A group of 57 (31 Spanish surveys, 26 English surveys) patients from the PNC constituted the intervention group. Prior to their clinic visit, participants engaged in a structured, one-on-one stroke education session with a member of the research team. This standardized intervention was designed to last approximately ten minutes while patients waited for their clinic visit. Patients were asked verbally if they had taken the pre-intervention survey and were excluded if they endorsed participating.

Immediately following the educational session, participants completed a post-intervention survey. This survey was identical to the one administered to the baseline (pre-intervention) group. The results from both groups were then compared to assess the efficacy of the educational intervention.

### Data Analytics and Statistical Analysis

Handwritten responses collected via standardized survey sheets were transcribed verbatim by research team members into a secure digital storage system. Responses submitted in Spanish were translated into English by native Spanish-speaking team members. All subsequent analyses were conducted in English.

Qualitative responses were analyzed using a directed content analysis approach,^25^ where a structured codebook was developed prior to data review to quantify the presence of “correct” knowledge within open-ended responses. Two independent coders, blinded to one another’s work, applied the codebook to all responses. Discrepancies between coders were resolved through consensus discussions, and an audit trail of coding decisions was maintained to ensure transparency and reliability.^26^

Coded data were processed in a Google Colab environment using Python to generate a non-binary matrix representing the presence of each code per response. This approach allowed for the quantification of coded themes and the calculation of percentage breakdowns for each categorical code. Matrices are attached as supplemental data.

While the analysis was primarily deductive, inductive methods were also employed to capture recurring themes not represented in the initial codebook.^27^ During double-coding, coders documented patterns or concepts that arose consistently outside of predefined categories. These emergent themes were reviewed during consensus discussions and summarized narratively as uncoded trends to complement the structured analysis. These uncoded trends were subsequently reported in the Results section alongside the quantitatively coded data.

For each language group, the proportions of responses coded under each category were compared between pre- and post-intervention groups using two-proportion z-tests. For each comparison, a z-score was calculated and used to derive a corresponding two-tailed p-value. Normality assumptions for the z-test were confirmed based on the sample size and distribution of responses. Statistical significance was defined as exact p <.05 in accordance with AMA reporting guidelines.

## Results

### Baseline Data

Demographic data shows a roughly even gender distribution (Table 1). Regarding race, there was a high rate of non-response, but among those who answered, 21% identified as White and 14.5% as Black. A majority identified as Hispanic or Latino and were of non-U.S. origin. The average participant age in the pre-intervention group was 48.8 years.

**Table 1.** Pre- and post-intervention participant demographics.

| **Demographic** | **Preintervention**<br>**Surveys**<br>**n=62 (%)** | **Postintervention**<br>**Surveys**<br>**n=57 (%)** |
| --- | --- | --- |
| **Gender** |  |  |
| Male | 29 (46.8) | 28 (49.1) |
| Female | 31 (50.0) | 29 (50.9) |
| Other | 1 (1.6) | 0 (0) |
| No response | 1 (1.6) | 0 (0) |
| **Race** |  |  |
| White | 13 (21.0) | 12 (21.1) |
| Black | 9 (14.5) | 8 (14.0) |
| Asian | 2 (3.2) | 4 (7.0) |
| Native American/Native Hawaiian/Pacific Islander | 4 (6.5) | 1 (1.8) |
| No response | 34 (54.8) | 32 (56.1) |
| **Ethnicity** |  |  |
| Non-hispanic or Latino | 18 (29.0) | 8 (14.0) |
| Hispanic or Latino | 37 (59.7) | 34 (59.6) |
| No response | 7 (11.3) | 15 (26.3) |
| **Country of Origin** |  |  |
| US origin | 20 (32.3) | 15 (26.3) |
| Non-US origin | 37 (59.7) | 41 (71.9) |
| No response | 5 (8.1) | 1 (1.8) |
| **Age (years)** | 48.8 | 42.4 |

Pre-intervention analysis demonstrated profound under-recognition of stroke warning signs across both language groups (Table 2). The lowest recognition was vision loss for both Spanish speakers (6.3%) and English speakers (6.7%). Recognition of all BEFAST/RÁPIDO stroke signs remained below 50% in both groups, except English recognition of speech problems (56.7%) and unilateral limb weakness (53.3%). Consequently, the full BEFAST/RÁPIDO model was incorporated into the educational intervention.

**Table 2.**
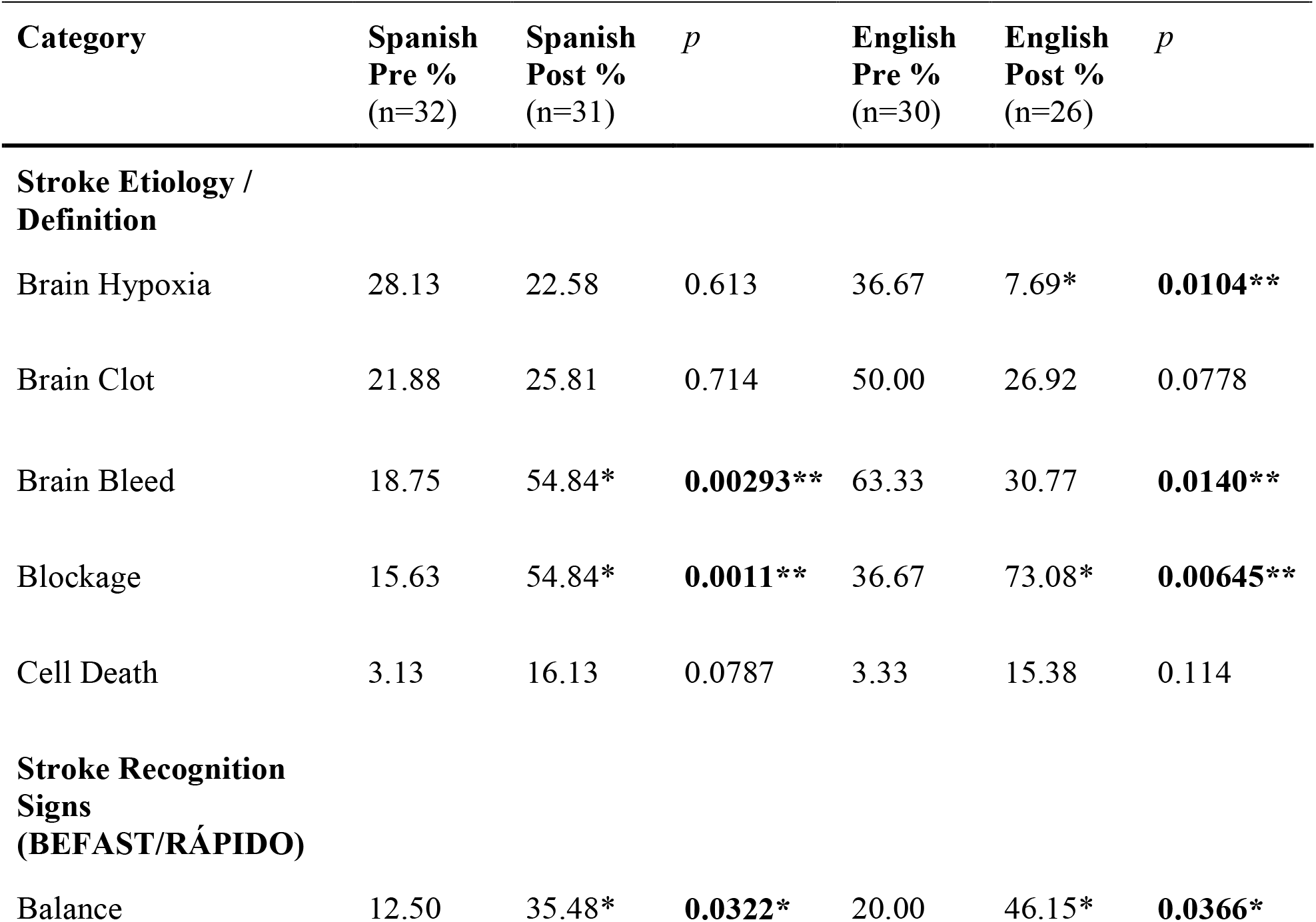
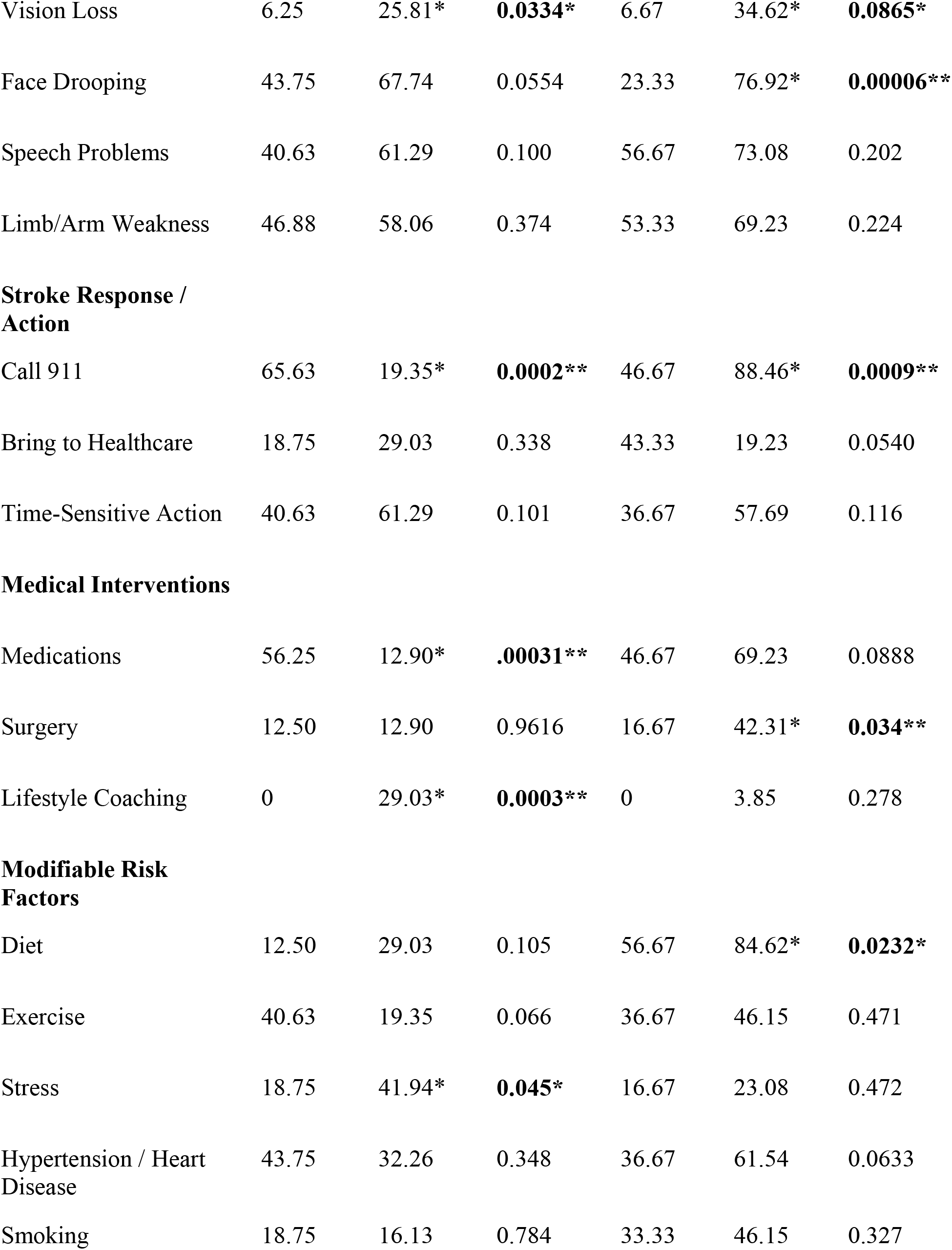
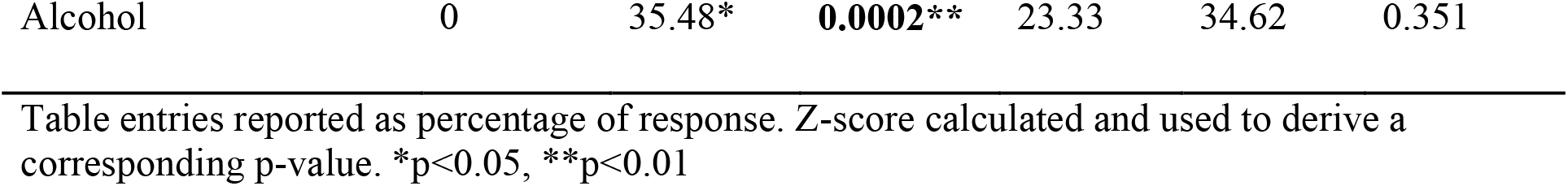
Stroke knowledge before and after an educational intervention, shown as percentages of participants who provided correct responses in Spanish-speaking and English-speaking groups.

| Category | Spanish<br>Pre %<br>(n=32) | Spanish<br>Post %<br>(n=31) | *p* | English<br>Pre %<br>(n=30) | English<br>Post %<br>(n=26) | *p* |
| --- | --- | --- | --- | --- | --- | --- |
| **Stroke Etiology /**<br>**Definition** |  |  |  |  |  |  |
| Brain Hypoxia | 28.13 | 22.58 | 0.613 | 36.67 | 7.69* | **0.0104\*\*** |
| Brain Clot | 21.88 | 25.81 | 0.714 | 50.00 | 26.92 | 0.0778 |
| Brain Bleed | 18.75 | 54.84* | **0.00293\*\*** | 63.33 | 30.77 | **0.0140\*\*** |
| Blockage | 15.63 | 54.84* | **0.0011\*\*** | 36.67 | 73.08* | **0.00645\*\*** |
| Cell Death | 3.13 | 16.13 | 0.0787 | 3.33 | 15.38 | 0.114 |
| **Stroke Recognition**<br>**Signs**<br>**(BEFAST/RÁPIDO)** |  |  |  |  |  |  |
| Balance | 12.50 | 35.48* | **0.0322\*** | 20.00 | 46.15* | **0.0366\*** |
| Vision Loss | 6.25 | 25.81* | **0.0334\*** | 6.67 | 34.62* | **0.0865\*** |
| Face Drooping | 43.75 | 67.74 | 0.0554 | 23.33 | 76.92* | **0.00006\*\*** |
| Speech Problems | 40.63 | 61.29 | 0.100 | 56.67 | 73.08 | 0.202 |
| Limb/Arm Weakness | 46.88 | 58.06 | 0.374 | 53.33 | 69.23 | 0.224 |
| **Stroke Response / Action** |  |  |  |  |  |  |
| Call 911 | 65.63 | 19.35* | **0.0002\*\*** | 46.67 | 88.46* | **0.0009\*\*** |
| Bring to Healthcare | 18.75 | 29.03 | 0.338 | 43.33 | 19.23 | 0.0540 |
| Time-Sensitive Action | 40.63 | 61.29 | 0.101 | 36.67 | 57.69 | 0.116 |
| **Medical Interventions** |  |  |  |  |  |  |
| Medications | 56.25 | 12.90* | **.00031\*\*** | 46.67 | 69.23 | 0.0888 |
| Surgery | 12.50 | 12.90 | 0.9616 | 16.67 | 42.31* | **0.034\*\*** |
| Lifestyle Coaching | 0 | 29.03* | **0.0003\*\*** | 0 | 3.85 | 0.278 |
| **Modifiable Risk Factors** |  |  |  |  |  |  |
| Diet | 12.50 | 29.03 | 0.105 | 56.67 | 84.62* | **0.0232\*** |
| Exercise | 40.63 | 19.35 | 0.066 | 36.67 | 46.15 | 0.471 |
| Stress | 18.75 | 41.94* | **0.045\*** | 16.67 | 23.08 | 0.472 |
| Hypertension / Heart Disease | 43.75 | 32.26 | 0.348 | 36.67 | 61.54 | 0.0633 |
| Smoking | 18.75 | 16.13 | 0.784 | 33.33 | 46.15 | 0.327 |
| Alcohol | 0 | 35.48* | **0.0002\*\*** | 23.33 | 34.62 | 0.351 |

Table entries reported as percentage of response. Z-score calculated and used to derive a corresponding p-value. \*p<0.05, \*\*p<0.01

Stroke-response awareness was also limited. Spanish participants most frequently failed to indicate bringing someone to healthcare (18.8%), while English participants least recognized stroke as time-sensitive (36.7%). Although 65.6% of Spanish speakers endorsed calling 911, fewer than half of English speakers did (46.7%). Educational emphasis was therefore placed on activating emergency medical services, urgency of time, and immediate presentation to healthcare.

Modifiable risk recognition was also low. The lowest response among Spanish-speakers was Alcohol as a risk factor (0%). The lowest response among English-speakers was Stress (16%). Smoking and exercise were less than 50% in both groups. All modifiable risk domains were included in the intervention, with special emphasis on the lowest-recognized item in each language group.

Baseline disparities demonstrated systematically lower Spanish recognition of bringing to healthcare (19% Spanish vs 43% English), diet (13% Spanish vs 57% English), and alcohol (0% vs 23%). Conversely, English participants had lower recognition of face drooping (23% English vs 44% Spanish) and bringing to healthcare (47% English vs 67% Spanish), illustrating heterogeneous gaps. These disparities were evaluated post-intervention for convergence.

### Post-Intervention Data

Demographic data, including gender, race, and ethnicity, was stable compared to the pre-intervention group. The percentage of participants not originally from the US increased from 59.7 to 71.9%. Participant age decreased to 42.4 from 48.8.

Stroke definition recognition remained low post-intervention, though select improvements were observed. Spanish participants showed significant gains in brain bleed (p < 0.01) and blockage (p < 0.01). English participants improved in blockage (p < 0.01) whereas brain hypoxia and brain bleed significantly decreased (p < 0.05). Brain hypoxia did not significantly change in Spanish participants, and cell death and brain clot did not significantly change in either language group.

Post-intervention, Spanish participants showed statistically significant improvement in recognition of balance (p < 0.05) and vision loss (p < 0.05). Face drooping, speech problems and limb/arm weakness did not significantly change in the Spanish speaking group. English participants significantly improved in balance (p < 0.05), vision loss (p < 0.01), and face drooping (p < 0.01), while speech problems and limb/arm weakness did not significantly change.

Among Spanish participants, Call 911 decreased significantly (p<0.01) and there was no change in Bring to Healthcare or Time-Sensitive Action. Among English speakers, calling 911 significantly improved (p < 0.01), but bring to healthcare and time sensitivity did not significantly change.

Knowledge of medical intervention for stroke response and prevention among Spanish speakers increased significantly for medications and lifestyle coaching (p < 0.01), but did not change significantly for surgery. Among English speakers, knowledge of surgery increased significantly (p < 0.05), but knowledge of medications and lifestyle coaching did not significantly change.

Modifiable risk factor knowledge among Spanish participants improved significantly for alcohol (p < 0.01) and stress (p < 0.05). Exercise, hypertension/heart disease, and smoking did not significantly change in Spanish participants. Among English participants, only diet (p < 0.05) improved significantly, while exercise, hypertension/heart disease, smoking, and heart disease did not significantly change.

In addition to the coded responses, an inductive theme emerged in the post-survey results. Among Spanish participants, several individuals reported fear of calling 911, even when they correctly identified stroke symptoms that could save a life. Out of 31 post-education Spanish surveys, coders independently noted this concern in n = 6 participants. No participants reported this theme in pre-surveys. These responses were summarized narratively, and all quotes are provided in Appendix D.

## Discussion

This educational intervention study demonstrates that providing targeted stroke education to English and Spanish speaking patients in a student-run free clinic can produce significant improvements in multiple domains of stroke knowledge. Compared to baseline survey results, participants showed measurable gains in stroke symptom recognition. The findings support the utility of the validated BEFAST/RÁPIDO symptom framework, which appeared to facilitate significant gains in symptom recognition. This is consistent with prior literature emphasizing the effectiveness of structured stroke symptom education in improving both recognition and response behaviors. ^28, 29, 30^

Improvements in stroke-response knowledge were limited. Among English speakers, only calling 911 demonstrated a statistically significant increase, whereas none of the stroke-response categories improved significantly among Spanish participants. Instead, Spanish participants’ reported likelihood of calling 911 significantly decreased from pre-intervention to post-intervention, despite significantly improved recognition of several stroke symptoms.

This apparent paradox may be partially explained by an inductive finding from the qualitative analysis: several Spanish participants expressed fear of contacting emergency services, even when recognizing that rapid response could be life-saving. Although the study did not identify the underlying causes of this fear, it highlights that even validated, structured educational tools may not fully overcome sociocultural or psychosocial barriers. Broader confounding variables such as concerns about immigration status, stressful political landscapes, distrust of healthcare systems, or prior negative experiences may influence emergency response behaviors in ways that are not addressed by symptom-focused education alone. These insights underscore the importance of culturally and contextually informed interventions when designing public health strategies for diverse populations.

Both statistically significant improvements and significant decreases in recognition across certain stroke definitions were observed, highlighting a nonuniform effect of the intervention on stroke definitions. Similarly, only a few improvements in knowledge of medical interventions and modifiable risk factors were observed. This may reflect the absence of a validated, structured framework for risk factor education analogous to BEFAST/RÁPIDO for symptoms. Participants showed selective gains in areas such as alcohol use, stress, diet, and hypertension awareness, suggesting that tailored emphasis on low-recognition risk factors can have some impact. Future interventions may benefit from integrating a structured, evidence-based risk-factor curriculum alongside symptom recognition training.

Notably, rates of improvement varied between the English and Spanish-speaking groups, even after validating materials with native speakers and utilizing in-person interpreters. This outcome highlights the persistent challenge and critical need for educational materials specifically designed for distinct linguistic and cultural groups. We plan to use these post-intervention findings to guide further refinement of our resources.

Overall, this study demonstrates that brief, linguistically and culturally tailored stroke education can be feasibly implemented within a student-run free clinic and meaningfully improve stroke warning sign recognition across underserved populations. However, stroke-response knowledge, particularly among Spanish speakers, appears constrained by confounding barriers, highlighting the need for future interventions to address fear-based reluctance to activate emergency services.

These findings support the potential of low-cost, scalable educational strategies to reduce stroke morbidity through earlier presentation and intervention, particularly in populations with language, literacy, or access barriers. Structured symptom frameworks such as BEFAST/RÁPIDO are effective for knowledge acquisition, but optimal outcomes may require additional attention to behavioral and cultural determinants of healthcare engagement.

This study is among the first to assess stroke literacy in a student-run free clinic. Educational materials were reviewed by stroke neurologists and translated into Spanish by native speakers to ensure accuracy and cultural appropriateness. The intervention is brief, accessible, and feasible for implementation in similar community-based settings. Quantitative coding of qualitative data allowed structured analysis while also capturing emergent themes, such as fear-based barriers to emergency response. To address potential limitations of the structured codebook, inductive methods were also applied to identify uncoded trends.

Limitations include a small sample size, single-site convenience sampling, and different individuals in the baseline and post-intervention groups, which may limit generalizability. Immediate post-intervention assessment prevents evaluation of long-term knowledge retention and health outcomes, which is the ultimate goal of an education program such as this. Environmental and sociocultural factors affecting stroke knowledge and response behaviors were not assessed or controlled. This introduces unforeseen confounding variables into the survey responses, which is true for any population-based study. Future work should evaluate knowledge retention and explore reinforcement strategies and effects on health behaviors.

## Conclusion

This study demonstrates that a brief, linguistically tailored stroke-education intervention can be successfully implemented in a student-run free clinic to address critical knowledge gaps among Spanish- and English-speaking underserved patients. The intervention produced meaningful improvements in recognition of BEFAST/RÁPIDO signs and select modifiable risk factors and offers a scalable, low-cost strategy to improve stroke literacy and support rapid action in high-risk populations. Notably, the findings highlight both the effectiveness of structured symptom education and the persistent sociocultural barriers, particularly among Spanish speakers, that may limit timely emergency response.

## Data Availability

All data produced in the present study are available upon reasonable request to the authors

## Sources of Funding

None.

## Disclosures

The authors have no conflicts of interest to disclose.

## Abbreviations

PNC: Phillips Neighborhood Clinic

## Notes

### Competing Interest Statement

The authors have declared no competing interest.

### Funding Statement

This study did not receive any funding

### Author Declarations

This study was determined exempt from review by the University of Minnesota Institutional Review Board (STUDY00023210).

